# Identifying temporal patterns in the progression of neurodegenerative disease using unsupervised clustering

**DOI:** 10.1101/2024.02.23.24303255

**Authors:** Alexia Giannoula, Audrey E. De Paepe, Ferran Sanz, Laura I. Furlong, Estela Camara

**Affiliations:** Epidemiology and Evaluation Department, Hospital del Mar Research Institute, Barcelona, Spain; Research Group on Integrative Biomedical Informatics (GRIB), Department of Medicine and Life Sciences (MELIS), Universitat Pompeu Fabra, Hospital del Mar Research Institute, Barcelona, Spain; Cognition and Brain Plasticity Unit, Institut d’Investigació Biomèdica de Bellvitge (IDIBELL), Barcelona, Spain; MedBioinformatics Solutions, Barcelona, Spain

**Keywords:** Precision medicine, longitudinal cohort analysis, multi-modal Real-World Data, patient stratification, unsupervised clustering, time analysis

## Abstract

**Objectives:** One of the principal goals of Precision Medicine is to stratify patients by accounting for individual variability. However, extracting meaningful information from Real-World Data, such as Electronic Health Records, still remains challenging due to methodological and computational issues.

**Materials & methods:** A Dynamic Time Warping-based unsupervised-clustering methodology is presented in this paper for the clustering of patient trajectories of multi-modal health data on the basis of shared temporal characteristics. Building on an earlier methodology, a new dimension of time-varying numerical clinical and imaging features (six in total) is incorporated, through an adapted cost-minimization algorithm for clustering on different, possibly overlapping, feature subsets. A cluster evaluation process is also implemented, by admitting two user-defined parameters (granularity threshold and feature contribution). The model disease chosen is Huntington’s disease (HD), characterized by progressive neurodegeneration.

**Results:** From a wide range of examined user-defined parameters, four case examples are highlighted to exemplify the combined effects of feature weights and granularity threshold in the stratification of HD trajectories in homogeneous clusters. For each identified cluster, polynomial fits that describe the temporal behavior of the assessed features are provided for an informative comparison, together with their averaged values.

**Discussion:** The proposed data-mining methodology permits the stratification of distinct time patterns of multi-modal health data in individuals that share a diagnosis or future diagnosis, employing user-customized criteria beyond the current clinical practice.

**Conclusions:** This work bears implications for better analysis of individual variability in disease progression, opening doors to personalized preventative, diagnostic and therapeutic strategies.

## Introduction

Precision medicine is an innovative approach that takes into account individual variability in genes, environment and lifestyle to understand diseases at a multifactorial level, with the aim of optimizing prevention, diagnostics and treatment of patient subgroups [1–3]. Such work has been empowered by the rapid expansion of Real-World Data (RWD), such as data stored within electronic healthcare records (EHRs), making precision medicine increasingly popular in recent years. In this way, individuals with a common diagnostic label may exhibit differences in terms of disease onset and response to treatment, characteristics that necessitate identification to maximize patient wellbeing. However, due to the challenges posed in extracting useful and actionable information from the large volumes of available data, there is an urgent need for the development of novel methodological approaches and computational tools to effectively stratify patients based on their relevant individual differences.

In this respect, the use of data-mining technology has introduced new prospects in the field of biomedical research [4–7], as it permits extracting a wealth of knowledge from health datasets that otherwise would remain hidden within patients’ clinical histories. In light of this, a growing number of studies has employed different algorithms to mine EHRs that mainly contain common clinical descriptors, such as diagnostic codes, human phenotypes, lab tests and other clinical features [8–12]. However, in the majority of these works, the temporal dimension is typically not taken into account or is considered in an insufficient way. It is only in the recent years that patterns of disease progression, referred to as trajectories, have been studied with the use of longitudinal patient cohorts [13–18]. These studies aimed to identify subgroups of patients exhibiting similar progression patterns by means of data-mining techniques for a variety of disease areas, including heart failure, diabetes mellitus or prostate cancer. This was achieved by monitoring the temporal dynamics of specific health outcomes, such as disease state or diagnostic codes.

In this study, we present an innovative data-mining methodological framework for the identification of distinct patient groups by analyzing the progression of a broad set of clinical and imaging features over time. This is achieved by significantly expanding and adapting the Dynamic Time Warping (DTW)-based unsupervised clustering methodology implemented in prior works [15,18,19], while incorporating a cost-minimization algorithm from [20]. The latter enables clustering on different and possibly overlapping subsets of the aforementioned features. Clustering trajectories based on a series of time-varying features constitutes an important contribution to the -so far-limited literature of data-mining techniques for longitudinal cohort analyses in the field of biomedicine.

To demonstrate the potential of the proposed methodology in elucidating the heterogeneity in patients’ clinical profiles, we employ Huntington’s disease (HD) as a model disease [21]. In brief, HD is an inherited neurodegenerative disease caused by a CAG repeat expansion in the *HTT* gene. Neurodegeneration in HD patients is progressive and debilitating. Brain atrophy initially targets the striatum, a deep brain structure whose subregions include the caudate, putamen and nucleus accumbens. [22–24]. The disease typically manifests in mid-adulthood with a characteristic triad of motor, cognitive and psychiatric features [25–27]. Gene-expansion carriers who will later develop the disease (e.g., premanifest individuals) can be identified prior to overt disease onset by elective genetic testing. Considering this and given its progressive nature, HD has previously been described as a model for neurodegeneration [28]. At the same time, HD is heterogeneous in clinical presentation and disease course, highlighting its potential for improved descriptions of disease progress and tailored therapeutic interventions. As such, given the variability in disease progression and the ability to identify susceptible individuals prior to overt disease onset, HD constitutes a suitable model to showcase the potential of the proposed methodology in revealing subgroups of patients with similar profiles of disease progression over time. Clinical evaluations describing the evolution of clinical features in all three domains (motor, cognitive and psychiatric) are considered, along with three target brain volumes derived from Magnetic Resonance Image (MRI): putamen, caudate and nucleus accumbens.

Overall, this study seeks to showcase the potential of the proposed methodology in longitudinal patient stratification through the use of multimodal clinical data sets extracted from EHRs. Its findings are expected to serve as a preliminary basis for better understanding the variability in disease profiles, in this instance exemplified in individuals with HD. Ultimately, the proposed methodology can be appropriately adapted and applied to any disease of interest and observational cohort, for which multidimensional metrics are available in the time axis.

## Methods

### Participants

The dataset consisted, initially, of forty-seven HD gene-expansion carriers (22 premanifest, 25 manifest). Three participants were excluded due to inability to undergo the MRI scan in the setting of claustrophobia and/or incomplete evaluation for an entire domain, resulting in 44 participants for the cluster analysis. Demographics of the whole sample are detailed in Supplementary Table S1. Premanifest individuals are clinically defined as those who have been genetically determined to have the *HTT* gene mutation but have not yet been formally diagnosed with HD by motor criteria. Hence, while not all participants displayed motor symptoms, each was a confirmed HD gene-expansion carrier at baseline (44.02 ± 3.05 CAG repeats). The standardized CAG-Age Product (CAP) score (CAP=100×age×(CAG–35.5)/627) was used as a measurement of HD state [29]. All participants were assessed with the Unified Huntington’s Disease Rating Scale total motor score (UHDRS-TMS) and total cognitive (UHDRS_cogscore) evaluation [30] (Supplementary Table S1). Neuropsychiatric evaluation was performed with the short-Problem Behavior Assessment (PBA-s) [30]. Details of clinical assessments are provided below. For each participant, there were a maximum total of six longitudinal visits, including baseline (mean number of 4.30 ± 1 assessments and mean inter-assessment duration of 14.0 ± 4.1 months). Thirty-six participants (81.8%) completed at least three visits and eight completed one or two visits. This resulted in 559 total clinical evaluations. Four premanifest participants converted to manifest over the course of the study.

### Clinical evaluation

The UHDRS-cogscore was employed to evaluate phonetic verbal fluency (F-A-S test) and psychomotor speed (Symbol Digit Modalities Test), as well as processing speed, attention and inhibitory control (word-reading, color-naming and interference components of the Stroop Test). To evaluate motor symptomatology, we employed the UHDRS-TMS, which quantifies dysarthria, chorea, dystonia, gait, postural stability and oculomotor function [31].

Neuropsychiatric features were evaluated using the PBA-s [32]. This semi-structured interview is administered in the presence of a knowledgeable informant and consists of eleven components: depressed mood, suicidal ideation, anxiety, irritability, angry or aggressive behavior, lack of initiative (apathy), preservative thinking/behavior, obsessive–compulsive behavior, paranoid thinking/behavior, hallucinations and disoriented thinking/behavior. Domains are calculated as the product of frequency × severity for each sign. A lower UHDRS-cogscore, in contrast to higher UHDRS-TMS and PBA-s scores, represents worse functioning. For this reason, total motor and psychiatric scores are coded negatively in the clustering algorithm.

All assessments were carried out by neurologists or neuropsychologists specializing in movement disorders. No participants reported previous history of neurological disorder other than HD. Data were anonymized, and the study was approved by the ethics committee of Bellvitge Hospital in accordance with the Helsinki Declaration of 1975 and all participants provided written informed consent.

### MRI acquisition and processing

MRI data were acquired with a 3T whole-body MRI scanner (Siemens Magnetom Trio; Hospital Clinic, Barcelona), using a 32-channel phased array head coil to procure structural T1-weighted images (magnetization-prepared rapid-acquisition gradient echo sequence), 208 sagittal slices, repetition time=1970ms, echo time=2.34ms, inversion time = 1050ms, flip angle = 9°, field of view = 256mm, 1mm isotropic voxel with no gap between slices.

Subcortical volumes for three brain structures, namely the caudate nucleus, putamen and nucleus accumbens, were estimated for each participant through an automated procedure for volumetric segmentation of the T1-weighted images using FreeSurfer (https://surfer.nmr.mgh.harvard.edu/). This parcellation allows the extraction of subcortical volumes that has previously been associated with neurodegenerative processes, including the total intracranial volume (TIV). TIV is used as a normalization factor, adjusting for head size differences. Volumes were averaged across the left and right for each anatomical region. For each participant, MRI data from a maximum of two longitudinal visits was obtained, over a period of approximately 18 months.

### Formation of HD trajectories

For an individual under consideration, the HD trajectory was defined as a finite time sequence of *T* chronologically ordered registries of clinical or MRI data as described above. A list of *K* = 6 clinical assessment scores was assigned at each time registry (Supplementary Table S2). These include the clinical evaluations (UHDRS-cogscore, UHDRS-TMS and PBA-s), as well as the three MRI-based volumes (caudate, putamen and nucleus accumbens), the latter of which are normalized by the TIV and then averaged over the corresponding left and right hemispheres. The aforementioned scores will be denoted hereafter as *features*, which form a *feature vector* at each time instant.

Subsequently, the HD trajectories corresponding to patients *i* = 1,2, …, *n*, can be written as:

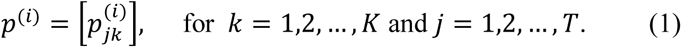

where the elements of the above temporal sequence represent registries at discrete times 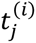 and *k* denotes the features assigned each time and *T* ≥ 1. Equation (1) can be expanded in two dimensions, yielding the following *K ×T* matrix for each HD patient trajectory:

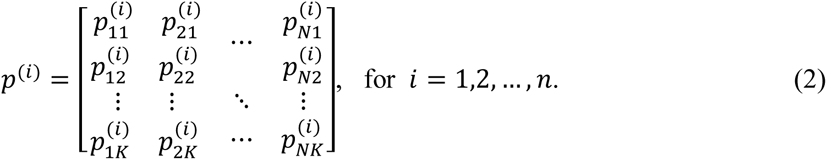

where *n* = 44.

Due to the variability in the number of longitudinal registries between all features, resampling method was applied on each feature and patient (method *resample* in MATLAB) in order to equalize the time length of all six feature vectors. This was achieved by upsampling each feature vector at *L_p_*/*L_o_* times the original sample rate, where *L_p_* denoted the maximum longitudinal length among all six feature vectors corresponding to a specific patient and *L_o_* the length of the original feature.

### DTW-based clustering of HD trajectories

Subsequently, the clinical trajectories of all HD patients were clustered in order to identify subgroups of patient profiles with shared temporal patterns. For this objective, the disease-trajectory clustering algorithm presented in [15,18] was adapted in order to incorporate not only the time, but also the feature dimension described earlier. In order to incorporate the time-varying feature vector into the clustering algorithm, the previous technique [20] for clustering objects on possibly overlapping subsets of features was modified.

In the adapted DTW-based clustering method proposed in this work, an HD trajectory was iteratively compared with the trajectories of the previously formed clusters by calculating their in-between distance. For two patients *i* and *q* with trajectories *p*^(*i*)^ and *p*^(*q*)^, respectively (described by equations (1) and (2)), the total distance between the two trajectories can be written as follows:

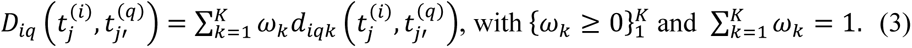

Time instants 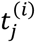 and 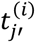 can be distinct in two patients, since the DTW algorithm permits variability in time scaling, length and in-between intervals, as noted in Introduction. The distances *d_iqk_*, of which the above equation is composed, refer to the different features under investigation and are given by:

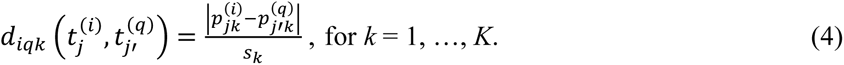

The denominator *s_k_* provides a scale for measuring “closeness” (spread) on each feature *k* over all patients of the cohort, i.e.:

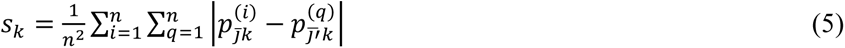

where the absolute distance is calculated, in a pair-wise manner for all patients of the cohort, at averaged times 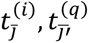 for each pair of patients.

The relative influence of each score 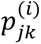 is regulated by its corresponding weight *ω_k_* in (3). Feature selection seeks to find an optimal weighting 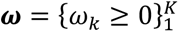 as part of the clustering problem by jointly minimizing the clustering criteria set in [20]. Although feature selection can be helpful, it attempts to form clusters based on the same subset of features. However, there may exist clusters whose groupings are formed based on different and possibly overlapping feature subsets. In this work, this kind of clustering was sought for the flexibility that it provides. For this reason, the generalized formula resulting from (3) was used, such that a separate score weighting 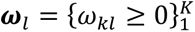 was defined for each cluster *C_l_*. As previously, the score weights satisfy 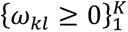 and 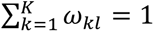

Finally, since the objective of this work was to extract clusters of patients that cluster simultaneously on subsets of scores (features), where each subset contains more than one score, the distance between two HD sequences *p*^(*i*)^ and *p*^(*q*)^, becomes:

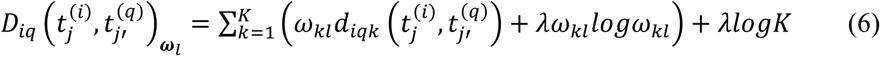

where the score weights are described by:

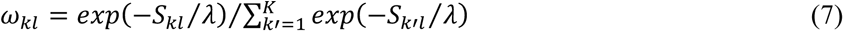

with 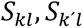 given by

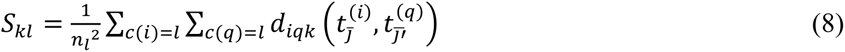

*S_kl_* is a measure of the dispersion (scale) of the data values on the *k*-th score for patients in the *l*-th cluster and *n_l_* defines the corresponding number of patients assigned to it.

At each iteration of the clustering algorithm, the score weights *ω_kl_* was calculated for each cluster and then the minimum averaged distance between each new HD trajectory and the trajectories already assigned to each cluster was sought. The algorithm terminated when there are no other HD trajectories to be considered.

### User-defined parameters and cluster evaluation

The DTW-based clustering algorithm presented above involved the selection of two user-defined parameters. First, the parameter *λ* seen in (6)-(7) controlled for the incentive for clustering on more features (*λ* ≥ 0), such that a lower or higher value of *λ* encouraged clusters on less or more scores, respectively. The score weights described by (7) put higher weight on features *k* with smaller dispersion (*S_kl_*) within each cluster *l*. Setting *λ* = 0 puts all weight on the feature with the smallest *S_kl_*, while *λ* = ∞ forces all features to contribute with equal weight to each cluster. Parameter *λ* will be denoted hereafter as *feature contribution parameter.* According to [20], there is no known mathematical formula to provide an optimal *λ* and therefore its value is typically selected based on empirical evaluation. In our clustering simulations, several values of *λ* were tested in order to assess different clustering configurations according to the degree of contribution for each score.

Furthermore, a granularity threshold *thres_gr_* was employed with the purpose of adjusting the clustering granularity. In this sense, lower values of *thres_gr_* increase cluster homogeneity at the cost of increasing the total number of clusters and vice versa. A compromise was generally sought to achieve sufficiently homogeneous clusters while avoiding excessive cluster fragmentation [15].

In order to optimize the selection of the above user-defined parameters, a cluster evaluation process was proposed that attempts to obtain a quantitative metric of the cluster homogeneity and, consequently, to assess the overall efficiency of the clustering algorithm. This involved the evaluation of different combinations of the granularity threshold and feature contribution parameter (*λ*) to identify clusters within HD trajectories. Subsequently, the total averaged distance over all extracted clusters was calculated, with lower values indicating increased homogeneity. In brief, for each combination of (*thres_gr_*, *λ*), the DTW-based unsupervised algorithm was applied and the cluster-based averaged distance was estimated for each identified cluster. This was achieved by averaging the pairwise distances between all HD trajectories assigned to the cluster under consideration based on the formula described in (6). The score weights *ω_kl_* used in that formula are the ones formed at the final step of the iterative clustering algorithm. The total averaged distance, corresponding to the examined pair of values (*thres_gr_*, *λ*), is then obtained by averaging across all extracted clusters.

Finally, the combination(s) of *thres_gr_* and *λ* that minimized the total averaged distance (thereby increasing cluster homogeneity), while controlling for the total number of extracted clusters was sought. In light of this, a compromise between the aforementioned conditions was needed in order to select the optimal user-defined parameters for the clustering algorithm. These parameters were empirically set to require (i) the total averaged distance to be < 30% of the maximum distance (resulting from all tested parameter values) and (ii) limiting the number of outlier clusters (those containing a single trajectory) to < 25% of the maximum number of extracted clusters.

### Linear fitting of the clustered HD trajectories

In the final step of the proposed methodological pipeline, an approximated linear characterization of the temporal patterns for the identified clusters was described. This was achieved by fitting each HD trajectory assigned to a cluster to a polynomial of degree 1 and averaging these over all the included trajectories. Consequently, seven approximated linear functions of time were obtained for each cluster, corresponding to (i) each feature trajectory individually (*y*_fit,*k*_, *k* = 1-6) and (ii) an averaged trajectory over all features (*y*_fit,all_).

## Results

The proposed DTW-based unsupervised algorithm was applied on the HD trajectories of *n* = 44 individuals using the following values for the feature contribution parameter *λ*: 0.01, 0.1, 0.2, 0.3, 0.4, 0.5, 0.6, 0.7, 0.8, 0.9, 1.0. With respect to the granularity threshold *thres_gr_*, a range of values between 0.5 and 8.0 was considered with intervals of 0.25. For each combination of *thres_gr_* and *λ*, the *averaged total distance* over all identified clusters was calculated as previously described along with the number of extracted clusters (Supplementary Fig. S1). Finally, nine pairs (*thres_gr_*, *λ*) were found to be optimal by fulfilling the requirements set in Methods, i.e.: (1.25, 0.01), (1.50, 0.01), (1.75, 0.01), (2.0, 0.1), (2.25, 0.01), (2.25, 0.1), (2.5, 0.01), (2.75, 0.01), (3.0, 0.01). Four case examples are highlighted below to demonstrate the effects of the aforementioned parameters on the identified clusters.

### Case 1: *thres_gr_* = 1. 50 and λ = 0.01

Case 1 demonstrated one of the optimal combinations that satisfied the conditions defined in the cluster evaluation process (see Methods), specifically *thres_gr_* = 1.50 and λ = 0.01. This resulted in a total of ten identified clusters as depicted in Fig. 1, with the total averaged distance of 0.66. Table 1 details averaged feature values, fitted polynomials and distribution of the final score weights (i.e., those reached at the final step of the iterative clustering algorithm). In the majority of clusters, only one or at most two features contributed to clustering, as was expected due to the very low selected value of λ.

**Fig. 1.**
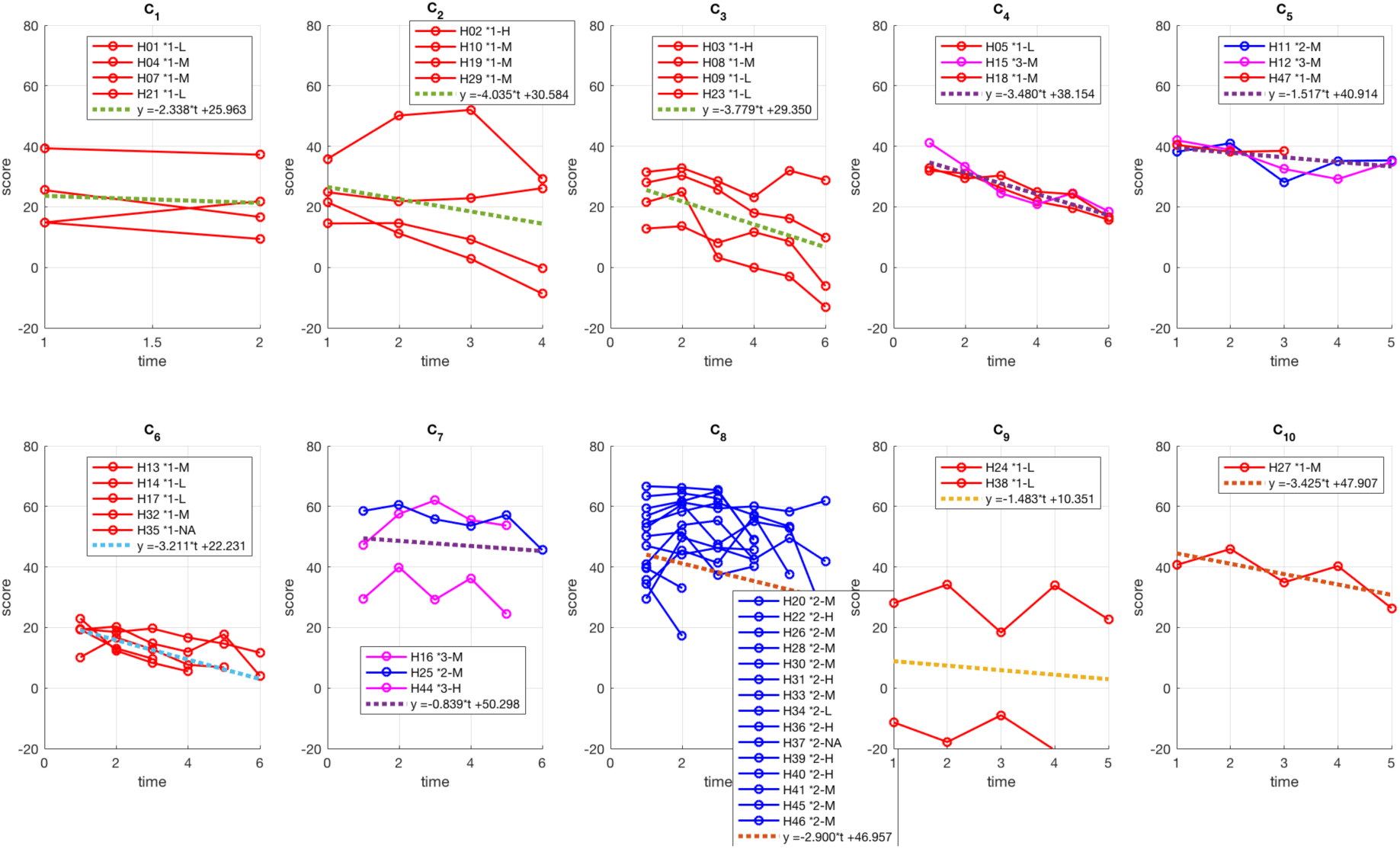
Extracted clusters of HD trajectories for Case 1 (*thres_gr_* = 1.50 and λ = 0.01). Red depicts manifest profiles, blue premanifest and pink those individuals that transitioned from premanifest to manifest over the course of assessment. The score on the y-axis is produced by averaging all six features at each time point. The averaged fitted polynomial of degree 1 is also shown (dotted lines) in each cluster.

**Table 1.** Case 1 individual *features* (*k* = 1-6) averaged over all patients and times in each cluster (*C_l_*, *l* = 1-10), for *thres_gr_* = 1.50 and λ = 0.01. The corresponding distribution of the score weights *ω_kl_* is also reported. For each cluster, the approximated polynomial of degree 1 is shown in the case of each feature trajectory separately (*y*_fit,*k*_, *k* = 1-6) and the averaged trajectory over all features (*y*_fit,all_).

| $C_l$ | 1 | 2 | 3 | 4 | 5 | 6 | 7 | 8 | 9 | 10 |
| --- | --- | --- | --- | --- | --- | --- | --- | --- | --- | --- |
| $k = 1$ | 171.74 | 169.23 | 149.96 | 188.81 | 242.69 | 135.31 | 306.26 | 299.40 | 128.78 | 237.79 |
| $k = 2$ | -24.68 | -26.63 | -33.75 | -17.17 | -11.69 | -42.21 | -3.21 | -0.64 | -43.98 | -8.60 |
| $k = 3$ | -12.32 | -19.63 | -19.46 | -15.81 | -10.82 | -10.77 | -18.57 | -14.00 | -46.60 | -3.40 |
| $k = 4$ | 1.13e-3 | 1.29e-3 | 1.01e-3 | 1.29e-3 | 1.59e-3 | 1.02e-3 | 1.54e-3 | 1.73e-3 | 9.32e-4 | 1.65e-3 |
| $k = 5$ | 1.46e-3 | 1.78e-3 | 1.30e-3 | 1.66e-3 | 1.97e-3 | 1.42e-3 | 2.02e-3 | 2.23e-3 | 1.17e-3 | 2.15e-3 |
| $k = 6$ | 1.37e-4 | 1.61e-4 | 1.15e-4 | 1.51e-4 | 1.65e-4 | 1.31e-4 | 2.21e-4 | 2.05e-4 | 1.13e-4 | 1.69e-4 |
| $\omega_{1l}$ | 0.000 | 0.000 | 0.000 | 1.000 | 1.000 | 0.999 | 0.000 | 0.000 | 0.000 | 0.1667 |
| $\omega_{2l}$ | 0.000 | 0.000 | 0.000 | 0.000 | 0.000 | 0.000 | 0.924 | 1.000 | 0.000 | 0.1667 |
| $\omega_{3l}$ | 0.000 | 0.000 | 0.000 | 0.000 | 0.000 | 0.001 | 0.000 | 0.000 | 0.000 | 0.1667 |
| $\omega_{4l}$ | 0.000 | 0.000 | 0.000 | 0.000 | 0.000 | 0.000 | 0.001 | 0.000 | 1.000 | 0.1667 |
| $\omega_{5l}$ | 1.000 | 1.000 | 0.469 | 0.000 | 0.000 | 0.000 | 0.076 | 0.000 | 0.000 | 0.1667 |
| $\omega_{6l}$ | 0.000 | 0.000 | 0.531 | 0.000 | 0.000 | 0.000 | 0.000 | 0.000 | 0.000 | 0.1667 |
| $y_{fit,all}$ | -2.3*t<br>+26.0 | -4.0*t<br>+30.6 | -3.8*t<br>+29.4 | -3.5*t<br>+38.2 | -1.5*t<br>+40.9 | -3.2*t<br>+22.2 | -0.8*t<br>+50.3 | -2.9*t<br>+54.5 | -1.5*t<br>+10.4 | -3.4*t<br>+47.9 |
| $y_{fit,1}$ | -10.5*t<br>+187.5 | -17.0*t<br>+211.7 | -15.6*t<br>+204.7 | -16.2*t<br>+245.5 | -8.3*t<br>+265.3 | -13.9*t<br>+172.2 | -7.9*t<br>+331.8 | -15.7*t<br>+290.6 | -4.7*t<br>+141.2 | -19.4*t<br>+296.1 |
| $y_{fit,2}$ | 1.1*t<br>-26.4 | -9.9*t<br>-2.0 | -5.5*t<br>-14.6 | -3.1*t<br>-6.4 | -2.2*t<br>-5.3 | -6.5*t<br>-26.0 | -1.5*t<br>+1.6 | -0.4*t<br>+0.2 | -4.5*t<br>-31.8 | -0.5*t<br>-7.1 |
| $y_{fit,3}$ | -4.6*t<br>-5.4 | 2.7*t<br>-26.2 | -1.6*t<br>-13.9 | -1.6*t<br>-10.1 | 1.4*t<br>-14.5 | 1.1*t<br>-12.8 | 4.4*t<br>-31.6 | -1.3*t<br>-9.1 | 0.3*t<br>-47.2 | -0.6*t<br>-1.6 |
| $y_{fit,4}$ | 0.6e-4*t<br>+1.0e-3 | -3.1e-4*t<br>+2.1e-3 | -2.1e-4*t<br>+1.7e-3 | -2.5e-4*t<br>+2.2e-3 | -3.6e-4*t<br>+2.6e-3 | -2.7e-4*t<br>+1.8e-3 | -3.1e-4*t<br>+2.5e-3 | -3.8e-4*t<br>+2.4e-3 | -2.9e-4*t<br>+1.7e-3 | -3.1e-4*t<br>+2.6e-3 |
| $y_{\text{fit},5}$ | -0.9e-4*t | -4.3e-4*t | -2.3e-4*t | -3.4e-4*t | -4.6e-4*t | -3.3e-4*t | -4.1e-4*t | -4.6e-4*t | -3.6e-4*t | -3.2e-4*t |
|  | +1.6e-3 | +2.9e-3 | +2.1e-3 | +2.8e-3 | +3.2e-3 | +2.4e-3 | +3.3e-3 | +2.9e-3 | +2.2e-3 | +3.1e-3 |
| $y_{\text{fit},6}$ | -0.9e-5*t | -4.7e-5*t | -2.2e-5*t | -3.2e-5*t | -3.8e-5*t | -3.0e-5*t | -4.7e-5*t | -4.0e-5*t | -3.5e-5*t | -4.5e-5*t |
|  | +1.5e-4 | +2.8e-4 | +1.9e-4 | +2.6e-4 | +2.7e-4 | +2.1e-4 | +3.7e-4 | +2.7e-4 | +2.1e-4 | +3.0e-4 |

A very clear distinction of individuals in the premanifest phase was observed, where the majority was assigned to cluster *C*_8_ based exclusively on the motor feature (*k* = 2). This cluster was distinguished by the presence of a very low level of motor symptoms, as evidenced by the fitted polynomial *y*_fit,2_ (Table 1). Furthermore, individuals in *C*_8_ (together with *C*_7_) presented the mildest cognitive and motor symptoms. In addition, the extent of brain atrophy in the caudate and putamen, represented by the MRI features, was the least in *C*_8_ compared to all ten clusters.

Conversely, manifest patients were further subdivided into distinct clusters according to the time characteristics of the corresponding associated feature vectors. Notably, two major subgroups (clusters *C*_1_ and *C*_2_) of manifest patients (group 1) were formed, clustered based on the atrophy of the MRI putamen volume (*k* = 5). Patients assigned to *C*_2_ demonstrated lower severity in all clinical scores and brain volumes (except psychiatric) at enrollment but deteriorated significantly faster than those in *C*_1_ Clusters *C*_4_-*C*_6_ were formed with the exclusive participation of the total cognitive score (*k* = 1) and specifically, patients in *C*_4_ exhibited one of the fastest rates of cognitive decline (*y_fit,_*_1_).

### Case 2: *thres_gr_* = 3. 75 and λ = 0.01

In Case 2, λ was kept consistent with Case 1 (λ = 0.01), but a higher granularity threshold was selected (*thres_gr_* = 3.75). Although these parameter values did not align with the list of empirically optimal combinations (see beginning of the Results section), they were selected to illustrate how certain parameters can result in a smaller number of extracted clusters, that is, coarser clustering at the cost of reduced homogeneity (see Methods). As such, the clustering algorithm converged into four clusters (Fig. 2), with an ensuing increase in total averaged distance (1.87 vs 0.66 in Case 1), as expected. Table 2 details averaged feature values, fitted polynomials and score weights.

**Fig. 2.**
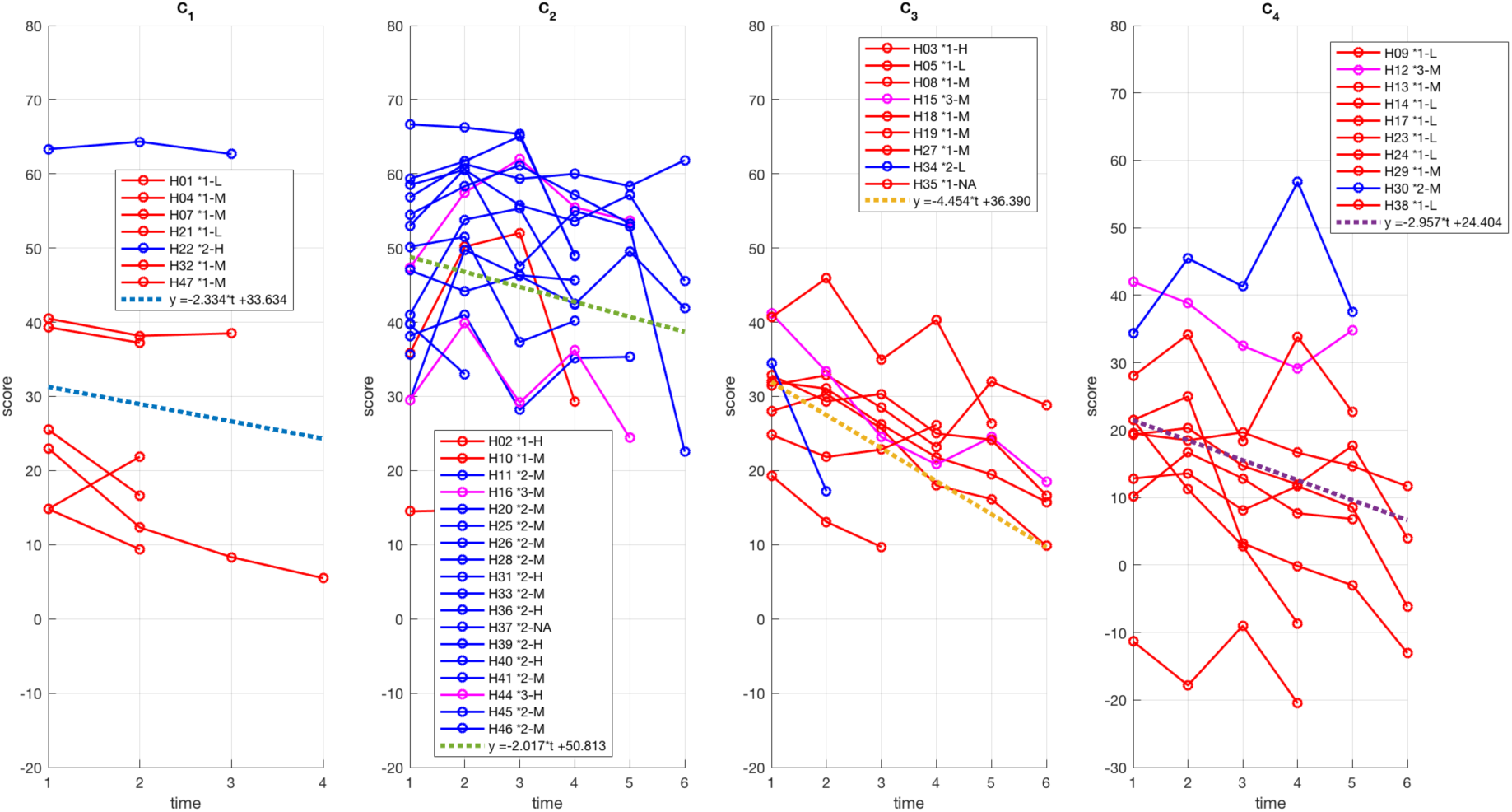
Extracted clusters of HD trajectories for Case 2 (*thres_gr_* = 3.75 and λ = 0.01). Red depicts manifest profiles, blue premanifest and pink those individuals that transitioned from premanifest to manifest over the course of assessment. The score on the y-axis is produced by averaging all six features at each time point. The averaged fitted polynomial of degree 1 is also shown (dotted lines) in each cluster.

**Table 2.**
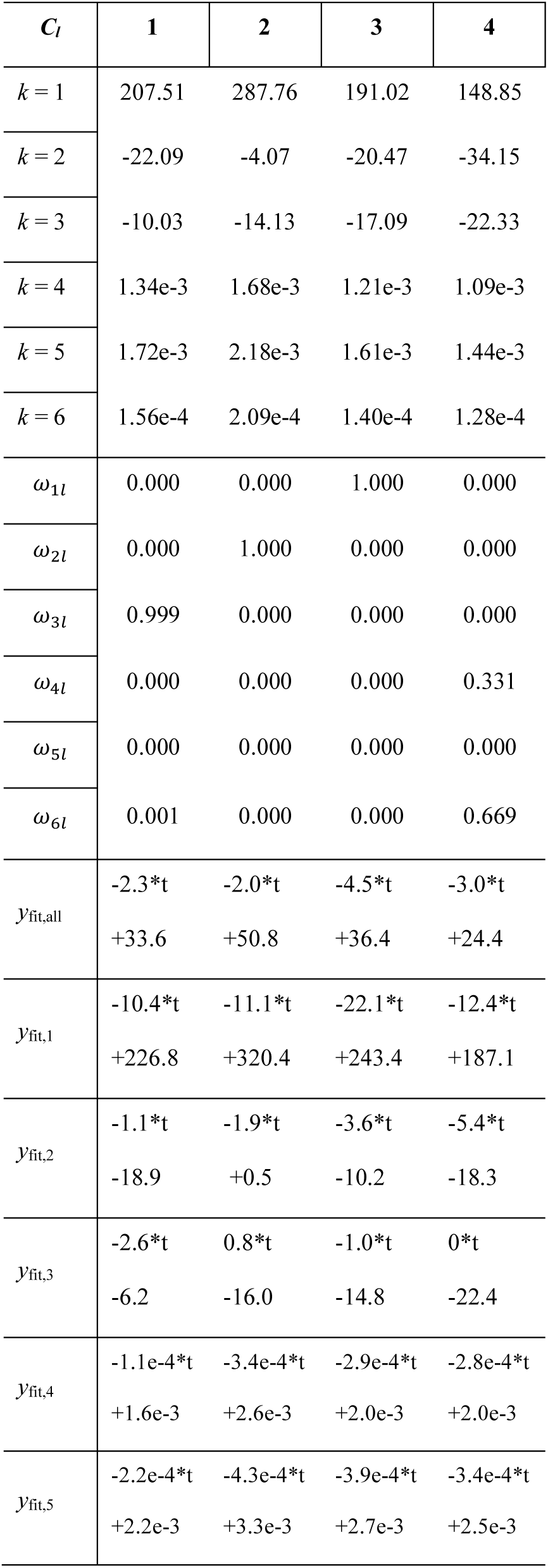
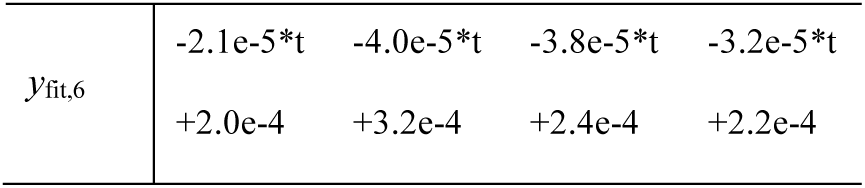
Case 2 individual *features* (*k* = 1-6) averaged over all patients and times in each cluster (*C_l_*, *l* = 1-4), for *thres_gr_* = 3.75 and λ = 0.01. The corresponding distribution of the score weights *ω_kl_* is also reported. For each cluster, the approximated polynomial of degree 1 is shown in the case of each feature trajectory separately (*y*_fit,*k*_, *k* = 1-6) and the averaged trajectory over all features (*y*_fit,all_).

| $C_l$ | 1 | 2 | 3 | 4 |
| --- | --- | --- | --- | --- |
| $k = 1$ | 207.51 | 287.76 | 191.02 | 148.85 |
| $k = 2$ | -22.09 | -4.07 | -20.47 | -34.15 |
| $k = 3$ | -10.03 | -14.13 | -17.09 | -22.33 |
| $k = 4$ | 1.34e-3 | 1.68e-3 | 1.21e-3 | 1.09e-3 |
| $k = 5$ | 1.72e-3 | 2.18e-3 | 1.61e-3 | 1.44e-3 |
| $k = 6$ | 1.56e-4 | 2.09e-4 | 1.40e-4 | 1.28e-4 |
| $\omega_{1l}$ | 0.000 | 0.000 | 1.000 | 0.000 |
| $\omega_{2l}$ | 0.000 | 1.000 | 0.000 | 0.000 |
| $\omega_{3l}$ | 0.999 | 0.000 | 0.000 | 0.000 |
| $\omega_{4l}$ | 0.000 | 0.000 | 0.000 | 0.331 |
| $\omega_{5l}$ | 0.000 | 0.000 | 0.000 | 0.000 |
| $\omega_{6l}$ | 0.001 | 0.000 | 0.000 | 0.669 |
| $y_{fit,all}$ | -2.3*t<br>+33.6 | -2.0*t<br>+50.8 | -4.5*t<br>+36.4 | -3.0*t<br>+24.4 |
| $y_{fit,1}$ | -10.4*t<br>+226.8 | -11.1*t<br>+320.4 | -22.1*t<br>+243.4 | -12.4*t<br>+187.1 |
| $y_{fit,2}$ | -1.1*t<br>-18.9 | -1.9*t<br>+0.5 | -3.6*t<br>-10.2 | -5.4*t<br>-18.3 |
| $y_{fit,3}$ | -2.6*t<br>-6.2 | 0.8*t<br>-16.0 | -1.0*t<br>-14.8 | 0*t<br>-22.4 |
| $y_{fit,4}$ | -1.1e-4*t<br>+1.6e-3 | -3.4e-4*t<br>+2.6e-3 | -2.9e-4*t<br>+2.0e-3 | -2.8e-4*t<br>+2.0e-3 |
| $y_{fit,5}$ | -2.2e-4*t<br>+2.2e-3 | -4.3e-4*t<br>+3.3e-3 | -3.9e-4*t<br>+2.7e-3 | -3.4e-4*t<br>+2.5e-3 |
| $y_{\text{fit},6}$ | -2.1e-5*t | -4.0e-5*t | -3.8e-5*t | -3.2e-5*t |
|  | +2.0e-4 | +3.2e-4 | +2.4e-4 | +2.2e-4 |

Similar to Case 1, Case 2 resulted again in one predominant premanifest cluster (*C*_2_), this time with three principally manifest clusters (*C*_1_, *C*_3_, *C*_4_). Specifically, the premanifest cluster *C*_2_ (similar to *C*_8_ in Case 1), was characterized by very low motor deficits with a relatively slow evolution over time (see *y_fit,_*_2_ in Table 2). On the other hand, the manifest cluster *C*_1_ was marked by the best (i.e., least severe) averaged total psychiatric symptoms but with the fastest deterioration in slope (*y_fit,_*_3_) of the psychiatric feature. Meanwhile, the manifest cluster *C*_3_ included mainly manifest patients undergoing the fastest decline of the total cognitive score (*y_fit,_*_1_) of all manifest clusters. Finally, *C*_4_ was formed based on brain atrophy of both the caudate and accumbens volumes, encompassing patients with the fastest decline in the total motor score (*y_fit,_*_2_) with stable evolution of the psychiatric feature (*y_fit,_*_3_) over time.

### Case 3: *thres_gr_* = 3. 75 and λ = 0.4

Case 3 exemplifies the effect of *λ* when patients are selectively grouped by a larger number of features. For this purpose, the granularity threshold was kept identical as in Case 2 (*thres_gr_* = 3.75) and *λ* was set equal to 0.4 (this combination of parameters is not listed as empirically optimal in the beginning of Results). Increasing *λ* permits a higher number of features to take part in the formation of each cluster. As a result, eight clusters were extracted (Fig. 3), and averaged feature values, fitted polynomials and score weights were produced (Table 3). The total averaged distance increased to 2.03 compared to Case 2, indicating a decrease in cluster homogeneity, as expected, since forming clusters with small dispersion in all or many features is not a practically optimal scenario (see Methods).

**Fig. 3.**
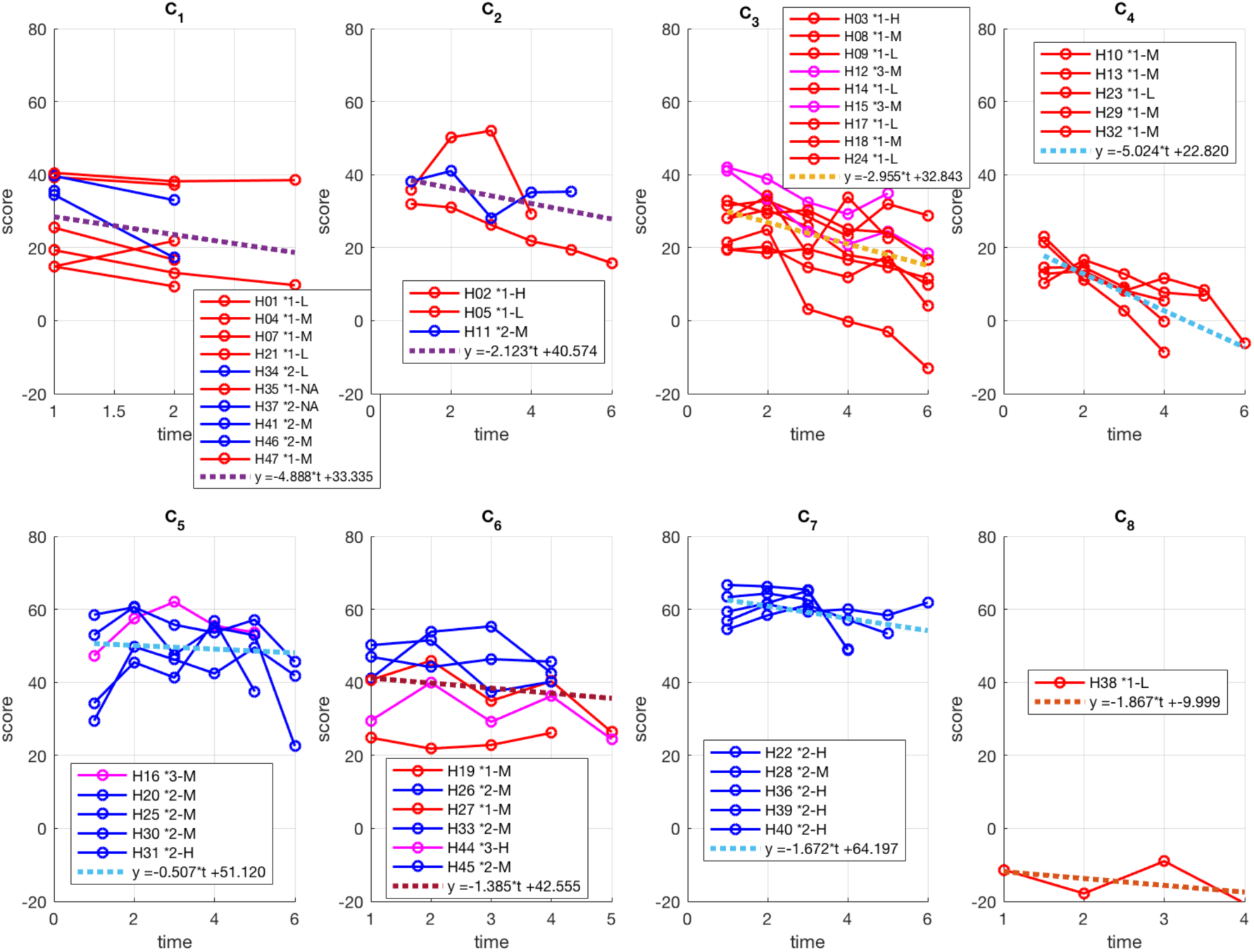
Extracted clusters of HD trajectories for Case 3 (*thres_gr_* = 3.75 and λ = 0.4). Red depicts manifest profiles, blue premanifest and pink those individuals that transitioned from premanifest to manifest over the course of assessment. The score on the y-axis is produced by averaging all six features at each time point. The averaged fitted polynomial of degree 1 is also shown (dotted lines) in each cluster.

**Table 3.** Case 3 individual *features* (*k* = 1-6) averaged over all patients and times in each cluster (*C_l_*, *l* = 1-8), for *thres_gr_* = 3.75 and λ = 0.4. The corresponding distribution of the score weights *ω_kl_* is also reported. For each cluster, the approximated polynomial of degree 1 is shown in the case of each feature trajectory separately (*y*_fit,*k*_, *k* = 1-6) and the averaged trajectory over all features (*y*_fit,all_).

| $C_l$ | 1 | 2 | 3 | 4 | 5 | 6 | 7 | 8 |
| --- | --- | --- | --- | --- | --- | --- | --- | --- |
| $k = 1$ | 203.46 | 231.16 | 177.85 | 116.03 | 312.84 | 251.23 | 363.51 | 41.75 |
| $k = 2$ | -16.99 | -7.25 | -27.01 | -43.46 | -1.47 | -4.61 | -0.78 | -62.75 |
| $k = 3$ | -18.10 | -20.34 | -14.69 | -15.61 | -17.01 | -14.48 | -2.08 | -67.00 |
| $k = 4$ | 1.39e-3 | 1.70e-3 | 1.09e-3 | 1.06e-3 | 1.62e-3 | 1.50e-3 | 1.86e-3 | 0.94e-3 |
| $k = 5$ | 1.88e-3 | 2.08e-3 | 1.46e-3 | 1.51e-3 | 2.02e-3 | 2.0e-3 | 2.27e-3 | 1.03e-3 |
| $k = 6$ | 1.80e-4 | 1.78e-4 | 1.32e-4 | 1.44e-4 | 1.89e-4 | 1.93e-4 | 1.88e-4 | 0.97e-4 |
| $\omega_{1l}$ | 0.421 | 0.130 | 0.140 | 0.297 | 0.140 | 0.130 | 0.248 | 0.1667 |
| $\omega_{2l}$ | 0.173 | 0.227 | 0.165 | 0.215 | 0.298 | 0.230 | 0.326 | 0.1667 |
| $\omega_{3l}$ | 0.161 | 0.209 | 0.061 | 0.122 | 0.108 | 0.050 | 0.270 | 0.1667 |
| $\omega_{4l}$ | 0.090 | 0.059 | 0.267 | 0.178 | 0.242 | 0.272 | 0.046 | 0.1667 |
| $\omega_{5l}$ | 0.084 | 0.069 | 0.180 | 0.125 | 0.112 | 0.266 | 0.026 | 0.1667 |
| $\omega_{6l}$ | 0.071 | 0.306 | 0.187 | 0.063 | 0.100 | 0.052 | 0.084 | 0.1667 |
| $y_{fit,all}$ | -4.9*t<br>+33.3 | -2.1*t<br>+40.6 | -3.0*t<br>+32.8 | -5.0*t<br>+22.8 | -0.5*t<br>+51.1 | -1.4*t<br>+42.6 | -1.7*t<br>+64.2 | -1.9*t<br>-10.0 |
| $y_{fit,1}$ | -24.3*t<br>+228.8 | -12.9*t<br>+270.0 | -12.6*t<br>+221.3 | -19.0*t<br>+166.6 | -8.8*t<br>+343.5 | -5.0*t<br>+266.1 | -10.0*t<br>+387.3 | -7.3*t<br>+60.0 |
| $y_{fit,2}$ | -1.2*t<br>-18.3 | -2.3*t<br>-0.7 | -4.1*t<br>-13.3 | -11.2*t<br>-14.1 | -0.6*t<br>+0.3 | -0.7*t<br>-2.6 | -0.5*t<br>+0.6 | -4.9*t<br>-50.5 |
| $y_{fit,3}$ | -3.8*t<br>-10.6 | 2.4*t<br>-25.8 | -1.1*t<br>-11.0 | 0.0*t<br>-15.6 | 6.4*t<br>-37.1 | -2.6*t<br>-8.2 | 0.5*t<br>-2.8 | 1.0*t<br>-69.5 |
| $y_{fit,4}$ | -1.0e-4*t<br>+1.4e-3 | -3.8e-4*t<br>+2.8e-3 | -2.4e-4*t<br>+1.9e-3 | -2.8e-4*t<br>+1.8e-3 | -3.6e-4*t<br>+2.8e-3 | -3.4e-4*t<br>+2.4e-3 | -4.2e-4*t<br>+3.0e-3 | -3.2e-4*t<br>+1.7e-3 |
| $y_{fit,5}$ | -2.3e-4*t<br>+2.0e-3 | -4.8e-4*t<br>+3.5e-3 | -3.0e-4*t<br>+2.5e-3 | -3.3e-4*t<br>+2.4e-3 | -4.5e-4*t<br>+3.5e-3 | -4.3e-4*t<br>+3.1e-3 | -4.9e-4*t<br>+3.6e-3 | -3.4e-4*t<br>+1.9e-3 |
| $y_{\text{fit},6}$ | -1.8e-5*t | -4.0e-5*t | -2.9e-5*t | -3.5e-5*t | -3.9e-5*t | -4.9e-5*t | -4.6e-5*t | -3.2e-5*t |
|  | +1.8e-4 | +3.0e-4 | +2.3e-4 | +2.4e-4 | +3.2e-4 | +3.2e-4 | +3.1e-4 | +1.8e-4 |

In Case 3, increasing the feature contribution parameter led to splitting of the major premanifest cluster of Case 2 (*C*_2_ in Fig. 2) into several smaller mainly premanifest or mixed clusters (e.g. *C*_1_, *C*_5_, *C*_6_, *C*_7_ in Fig. 3) and the rearrangement of the three principal manifest clusters of Case 2. In particular, the two purely premanifest clusters *C*_5_ and *C*_7_ contained the least severe cases of all with similar evolution over time in basically all clinical and brain-volume features except for the psychiatric domain. In this respect, patients in *C*_5_ exhibited a significantly worse psychiatric score than *C*_7_ at enrollment that improved relatively quickly with time, while patients in *C*_7_ remained relatively stable at higher (less severe) levels (*y_fit,_*_3_ in Table 3). At the same time, Case 3 demonstrated two purely manifest clusters (*C*_3_ and *C*_4_), where manifest patients in *C*_4_ were overall more severe than *C*_3_ and deteriorated faster in all clinical and brain-volume features, except for the psychiatric domain that was stable through time (*y_fit,_*_3_).

### Case 4: *thres_gr_* = 6. 0 and λ = 0.4

In Case 4, the granularity threshold *thres_gr_* was further increased to 6.0, while *λ* was maintained at 0.4 (this combination of parameters is also not listed as empirically optimal in the beginning of Results). This resulted in the coarsest possible separation, dividing patients into a manifest (*C*_1_) and premanifest (*C*_2_) cluster (Fig. 4). The resulting total averaged distance was the highest compared to all previous case examples, reaching 3.75. Furthermore, although all six features contributed to the formation of the two clusters (see score weights *ω_kl_* in Table 4), the most notable differences were seen in the total motor deficits (*k* = 2), which was significantly lower and deteriorated at a slower rate in *C*_2_ than *C*_1_. Interestingly, the decline in the total cognitive score was similar in both clusters (*y*_fit,1_), and the brain atrophy represented by the three MRI features was faster in premanifest than in the manifest group, albeit with higher values (i.e., less atrophy) at the time of enrollment (*y*_fit,4,_ *y*_fit,5_, *y*_fit,6_), as expected.

**Fig. 4.**
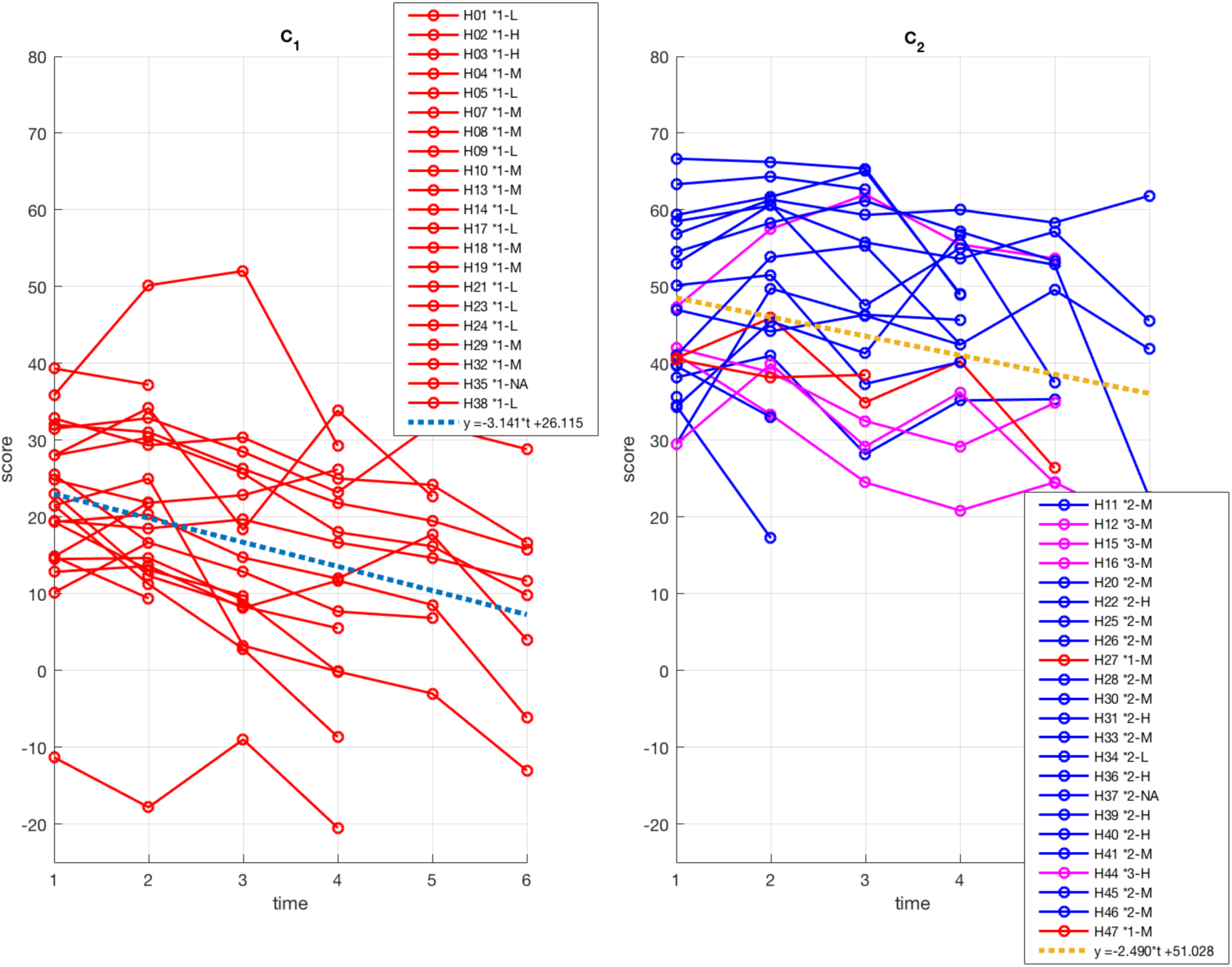
Extracted clusters of HD trajectories for Case 4 (*thres_gr_* = 6.0 and λ = 0.4). Red depicts manifest profiles, blue premanifest and pink those individuals that transitioned from premanifest to manifest over the course of assessment. The score on the y-axis is produced by averaging all six features at each time point. The averaged fitted polynomial of degree 1 is also shown (dotted lines) in each cluster.

**Table 4.**
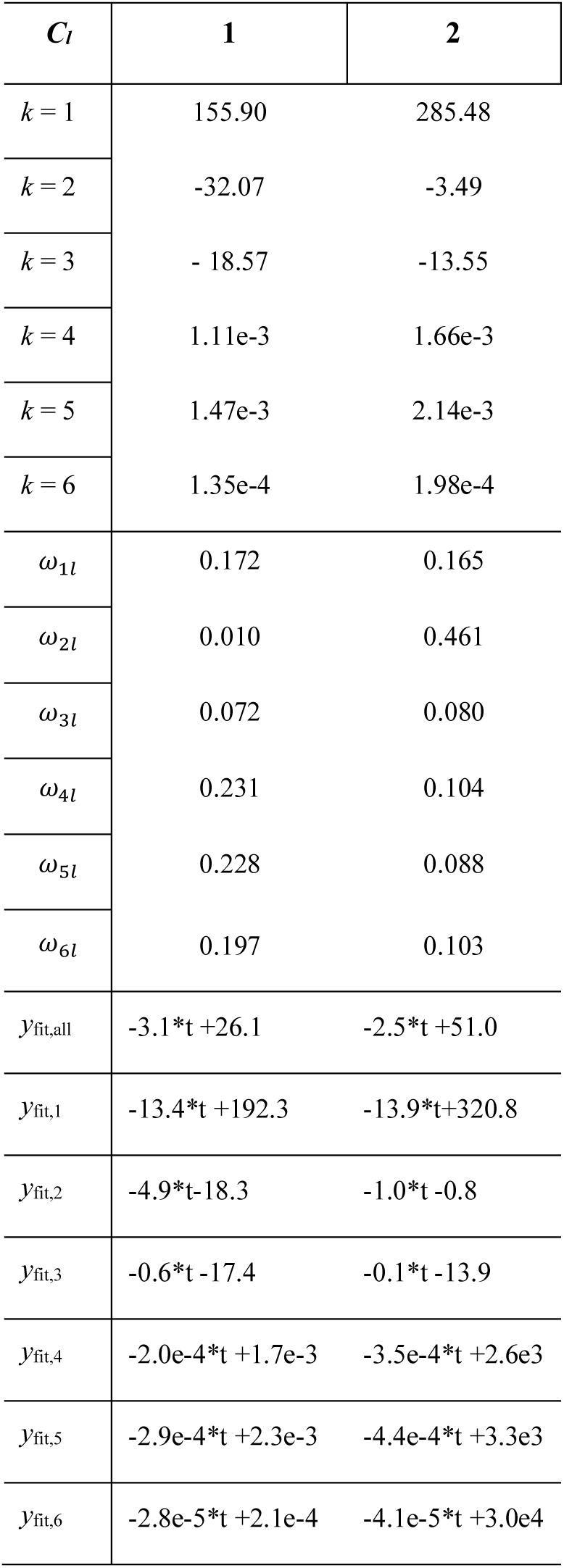
Case 4 individual *features* (*k* = 1-6) averaged over all patients and times in each cluster (*C_l_*, *l* = 1,2), for *thres_gr_* = 6.0 and λ = 0.4. The corresponding distribution of the score weights *ω_kl_* is also reported. For each cluster, the approximated polynomial of degree 1 is shown in the case of each feature trajectory separately (*y*_fit,*k*_, *k* = 1-6) and the averaged trajectory over all features (*y*_fit,all_).

| $C_l$ | 1 | 2 |
| --- | --- | --- |
| $k = 1$ | 155.90 | 285.48 |
| $k = 2$ | -32.07 | -3.49 |
| $k = 3$ | - 18.57 | -13.55 |
| $k = 4$ | 1.11e-3 | 1.66e-3 |
| $k = 5$ | 1.47e-3 | 2.14e-3 |
| $k = 6$ | 1.35e-4 | 1.98e-4 |
| $\omega_{1l}$ | 0.172 | 0.165 |
| $\omega_{2l}$ | 0.010 | 0.461 |
| $\omega_{3l}$ | 0.072 | 0.080 |
| $\omega_{4l}$ | 0.231 | 0.104 |
| $\omega_{5l}$ | 0.228 | 0.088 |
| $\omega_{6l}$ | 0.197 | 0.103 |
| $y_{fit,all}$ | $-3.1*t+26.1$ | $-2.5*t+51.0$ |
| $y_{fit,1}$ | $-13.4*t+192.3$ | $-13.9*t+320.8$ |
| $y_{fit,2}$ | $-4.9*t-18.3$ | $-1.0*t-0.8$ |
| $y_{fit,3}$ | $-0.6*t-17.4$ | $-0.1*t-13.9$ |
| $y_{fit,4}$ | $-2.0e-4*t+1.7e-3$ | $-3.5e-4*t+2.6e3$ |
| $y_{fit,5}$ | $-2.9e-4*t+2.3e-3$ | $-4.4e-4*t+3.3e3$ |
| $y_{fit,6}$ | $-2.8e-5*t+2.1e-4$ | $-4.1e-5*t+3.0e4$ |

## Discussion and Conclusions

This study presents an innovative data-mining methodological pipeline designed to cluster patients into subgroups that share similar profiles of time-varying multimodal clinical and imaging features. To demonstrate the potential of the current methodology in elucidating individual heterogeneity hidden beneath a common diagnosis, HD was employed as a model. By analyzing the severity and progression of six target clinical and imaging features in HD patients over time, this work serves as a proof-of-concept in the stratification of patients with a current or impending diagnostic label into groups with similar disease trajectories. The present approach expands on earlier unsupervised-clustering methodology [15,18,19] by incorporating a new dimension of numerical features through an adapted cost-minimization algorithm for clustering on different, possibly overlapping, feature subsets. In this manner, a valuable contribution is provided to the currently limited literature on data-mining techniques for longitudinal cohort analyses in biomedicine.

Methodologically, the study explores various user-defined parameters, such as the granularity threshold and feature contribution parameter (λ), showcasing results obtained under quantitatively optimized clustering parameters while allowing flexibility in user-defined parameters. Specifically, Case 1 was defined by optimum parameters (granularity threshold and feature contribution), as marked by the smallest total averaged distance, producing ten clusters (Fig. 1). Case 2 shared the same feature contribution parameter as Case 1, but with a higher granularity threshold, in order to produce coarser clustering (i.e., fewer clusters) at the cost of reduced homogeneity. In this case, patient profiles previously assigned to ten clusters (Fig. 1) were merged and/or re-organized to finally form four clusters (Fig. 2). For example, the premanifest cluster *C*_2_ of Case 2 (Fig. 2) included the majority of the premanifest patients from *C*_8_ of Case 1 (Fig. 1), as well as five additional patients (from different clusters of Fig. 1), all exhibiting mild motor symptoms.

Case 3 had an identical granularity threshold as in Case 2, with an increased feature contribution parameter, in order to demonstrate the result of permitting a higher number of features to take part in the formation of each cluster. As a result, eight clusters were extracted (Fig. 3) with a resulting decrease in cluster homogeneity, since forming clusters with small dispersion in all or many features is not a practically optimal scenario (see Methods). Finally, Case 4 was defined by an increase in the granularity threshold, while maintaining the feature contribution parameter the same as Case 3. This resulted in the coarsest possible separation, dividing patients into a manifest (*C*_1_) and premanifest (*C*_2_) cluster (Fig. 4). In addition, the resulting total averaged distance was the highest compared to all previous case examples, demonstrating that clusters were the least homogeneous of the case series. Of note, this coarse division between premanifest and manifest is that which is currently utilized in the clinical context, based solely on the motor score and a diagnostic confidence score determined by the clinician. Ultimately, this intra-diagnostic heterogeneity further underscores the potential for the current methodology to further refine diagnostic criteria and prognostic predictions.

In summary, the proposed data-mining methodology illustrates how temporal differences in the disease progression of patients diagnosed with HD can be identified and evaluated, in this case leveraging both clinical data and imaging biomarkers. Furthermore, the methodology allows for user-defined flexibility that allows exploration of data through different lenses, while also promoting a quantitative metric, such as the cluster-evaluation process, to assess the optimization of clustering parameters. While the main focus of the present study was not to interpret all extracted patterns nor their causality, this work establishes a framework for better understanding the observed heterogeneity in the disease progression of patients. It should be noted that inherent to the nature of the health dataset used, there may be incomplete data and errors in the clinical time registries and/or dates and thus, the results should be interpreted accordingly.

Considering individual variability constitutes a basic principle of precision medicine, which strives to tailor preventative, diagnostic and therapeutic strategies to the level of a single patient or subgroup of patients. In this manner, future directions could investigate the elaboration of personalized therapeutic management or inclusion of patients with similar features into clinical trials, even in early stages of the disease and across disease categories (e.g., premanifest vs. manifest, pre-diabetic vs. diabetic, Stage I vs. Stage II breast cancer, or depression in bipolar II vs. major depressive disorder).

In conclusion, this research introduces a novel data-mining methodological framework that integrates temporal characteristics of a set of multimodal clinical and imaging features of patients. In this way, this work enables the identification of distinct longitudinal disease profiles within big datasets such as EHRs in the context of RWD, further promoting personalized medicine for the improvement of human health. The flexible nature of the methodology allows for future expansions that incorporate additional longitudinal patient information, such as prescription drugs or other imaging and laboratory data.

## Acknowledgements

The authors are grateful to the patients and their families for their participation in this project. We would also like to thank Dr. Saül Martinez-Horta, Dra. Andrea Horta-Barba Dr. Jesús Pérez Pérez, Dr. Jaime Kulisevksy, Pilar Sanchez, Dr. Esteban Muñoz, Celia Mareca, Dr. Ruiz-Idiago, Dra. Matilde Calopa, Nadia Rodriguez-Dechichá, Irene Vaquer, Yemila Plana, Dra. Matilde Calopa and Dra. Clara Garcia-Gorro for help with clinical evaluation and data collection.

## Author Contributions

A. G. designed the methodology, performed the simulations and analysis of data and drafted the manuscript. A. E. D. helped conceive the project idea and drafted parts of the paper relevant to Huntington’s disease, the study participants and their clinical/MRI evaluation. She also revised the paper and helped in interpreting the results. L. F. and F. S. revised and approved the manuscript. E. C. revised the manuscript, helped in interpreting the results and supervised the work.

## Ethics Approval

All data contained in the cohort is anonymized. The study was approved by the ethics committee of Bellvitge Hospital in accordance with the Helsinki Declaration of 1975 and all participants provided written informed consent.

## Data Availability

The data underlying this article cannot be shared publicly for the privacy of individuals that participated in the study (ethics approval by Bellvitge Hospital, Spain, in accordance with the Helsinki Declaration of 1975). The data will be shared on reasonable request to the corresponding author.

