## Supplementary file for "Identifying temporal patterns in the progression of neurodegenerative disease using unsupervised clustering"

**Supplementary Table S1.** Sociodemographic, clinical, and neuroimaging characteristics of study participants (initial value).

| | **Premanifest** | **Transition** | **Manifest** | **HD gene-carriers** |
|---|---|---|---|---|
| *n* | 17 | 4 | 23 | 44 |
| Sex (f/m) | 13 / 4 | 4 / 0 | 14 / 9 | 31 / 13 |
| Age (years) | 36.12 ± 9.75 | 34.00 ± 2.94 | 49.22 ± 9.24 | 42.77 ± 11.25 |
| Education (years) | 14.06 ± 4.12 | 12.75 ± 2.99 | 11.87 ± 3.98 | 12.80 ± 4.02 |
| CAG | 43.71 ± 2.64 | 44.00 ± 2.83 | 44.26 ± 3.45 | 44.02 ± 3.05 |
| CAP | 45.16 ± 13.39 | 45.14 ± 10.65 | 65.00 ± 17.88 | 55.53 ± 18.40 |
| UHDRS-TMS | 0.35 ± 0.61 | 5.00 ± 4.24 | 23.00 ± 13.49 | 12.61 ± 14.73 |
| UHDRS-cogscore | 303.29 ± 54.18 | 291.67 ± 56.66 | 181.96 ± 54.74 | 237.34 ± 79.65 |
| PBA-s total | 19.88 ± 23.26 | 30.50 ± 35.24 | 18.35 ± 15.83 | 20.05 ± 20.68 |
| Caudate volume* | 2822.82 ± 465.54 | 2099.44 ± 256.20 | 1904.82 ± 377.14 | 2277.19 ± 593.97 |
| Putamen volume* | 3594.12 ± 631.88 | 2796.33 ± 323.04 | 2471.80 ± 455.40 | 2934.93 ± 741.79 |
| Nucleus accumbens volume* | 325.76 ± 67.41 | 306.76 ± 49.35 | 231.40 ± 53.03 | 274.71 ± 73.68 |

Data presented as *mean±standard deviation*. Premanifest and manifest grouped based on Unified Huntington's Disease Rating Scale diagnostic confidence score for motor abnormalities at first visit (Huntington Study Group, 1996). Transition indicates premanifest individuals that converted to manifest over the course of the study.

*Averaged MRI volume across right and left hemispheres for each participant.

CAG=length of cytosine-adenine-guanine base length repeats of the mutated allele; CAP=standardized CAG-age product f=females; HD=Huntington's Disease; m=males; *n*=number of participants; PBA-s=short-Problem Behavior Assessment; UHDRS-cogscore=Unified Huntington's Disease Rating Scale total cognitive; UHDRS-TMS=Unified Huntington's Disease Rating Scale total motor score.

**Supplementary Table S2.** Description of *features* that form the feature vector in the clustering of HD trajectories.

| *feature* | **Description** |
|---|---|
| $k = 1$ | Total cognitive score (UHDRS-cogscore) |
| $k = 2$ | Total motor score (UHDRS-TMS) |
| $k = 3$ | Total psychiatric score (PBA-s) |
| $k = 4$ | Caudate volume |
| $k = 5$ | Putamen volume |
| $k = 6$ | Nucleus accumbens volume |

*Averaged MRI volume across right and left hemispheres for each participant.
HD=Huntington's Disease; MRI=Magnetic Resonance Imaging; PBA-s=short-Problem Behavior Assessment; UHDRS-cogscore=Unified Huntington's Disease Rating Scale total cognitive score; UHDRS-TMS=Unified Huntington's Disease Rating Scale total motor score.

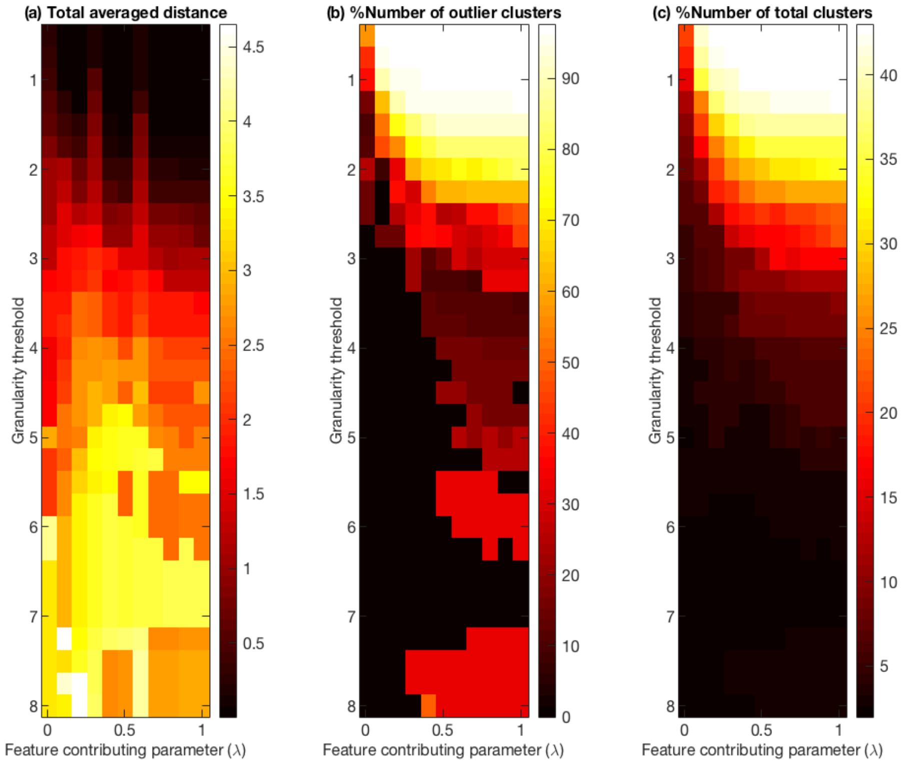


**Supplementary Fig. S1. Optimization parameters of the DTW-based unsupervised algorithm.** Heatmaps of (a) total averaged distance, (b) percent (%) number of outlier (single-trajectory) clusters and (c) number of total clusters extracted for different values of the granularity threshold ($thres_{gr}$) and feature contribution parameter ($\lambda$).
